# The Landscape of RAD51D in Chinese Ovarian Cancer Patients: Prevalence, Correlation with HRD Score, and Correlation with Efficacy

**DOI:** 10.1101/2024.03.19.24304522

**Authors:** Tianqi Sun, Mengpei Zhang, Xinyun Xu, Yiming Liang, Jinghong Chen, Qingli Li, Jing Zeng, Zhe Li, Yu Dong, Rutie Yin

## Abstract

Human genome is continuously threatened by endogenous and exogeneous sources of DNA damage, DNA damage repair (DDR) mechanisms are vital for genome instability and cancer prevention. Homologous recombination repair (HRR) is one of the most important DDR mechanisms because of its high-fidelity. BRCA1/2 have long been the best characterized HRR genes, but other non-BRCA1/2 HRR genes should not be overlooked. In this study, we illustrate that RAD51D is an important non-BRCA1/2 HRR gene in Chinese ovarian cancer patients, from perspectives including prevalence (2.9% patients mutated in RAD51D, 72.7% of RAD51D mutations were bi-allelic loss-of-function [BILOF]), correlation with HRD score (P=0.295 for RAD51D mutation, P=0.087 for RAD51D BILOF), and correlation with efficacy (progression-free survival: HR=0.685 P=0.069 for RAD51D mutation, HR=0.571 P=0.054 for RAD51D BILOF).

## 1. Introduction

The estimated number of new cases of ovarian cancer in China in 2022 was 61100, with an age-standardized incidence rate of 5.68 per 100000, ranked the 9th place in females (Han et al., 2024). The estimated number of new cases of ovarian cancer worldwide in 2020 was 313959, with an age-standardized incidence rate of 6.6 per 100000, ranked the 8th place for incidence rate in females (Cabasag et al., 2022). The cumulative risk to age 75 for developing ovarian cancer was estimated to be 0.73% in 2020 worldwide (Ferlay et al., 2021). The age-standardized incidence rate in 2020 was estimated to be 8.1 per 100000 in North America, while that in Eastern Asia was estimated to be 5.7 per 100000 (Cabasag et al., 2022).

The estimated number of deaths from ovarian cancer in China in 2022 was 32600, with an age-standardized mortality rate of 2.64 per 100000, ranked the 9th place in females (Han et al., 2024). The estimated number of deaths from ovarian cancer worldwide in 2020 was 207252, with an age-standardized mortality rate of 4.2 per 100000, ranked the 8th place for mortality rate in females (Cabasag et al., 2022). The cumulative risk to age 75 for death from ovarian cancer was estimated to be 0.49% in 2020 worldwide (Ferlay et al., 2021). The age-standardized mortality rate in 2020 was estimated to be 4.1 per 100000 in North America, while that in Eastern Asia was estimated to be 3.3 per 100000 (Cabasag et al., 2022).

Most epithelial serous ovarian cancer patients were diagnosed at stage III (51%) and stage IV (29%), and the 5-year survival rates were 42% and 26% for stage III and IV patients, respectively, reflecting the aggressive nature of serous ovarian cancer (Torre et al., 2018). Most non-serous epithelial ovarian cancers, including endometroid, mucinous, and clear cell carcinomas, were diagnosed at stage I (59%, 64%, and 58%, respectively), the 5-year survival rates for stage I endometroid, mucinous, and clear cell carcinoma patients were 95%, 92%, and 85%, respectively (Torre et al., 2018). Ovarian cancer diagnosed at local stage had a 5-year survival rate of 93%, therefore early detection of ovarian cancer has long been a research priority (Torre et al., 2018).

It was estimated that approximately 20%-25% women harbored inherited germline mutation that predisposes them to ovarian cancer (Kanchi et al., 2014; Walsh et al., 2011; Weissman et al., 2012). Approximately 31% of ovarian cancer patients had germline or somatic mutations in one of the 13 homologous recombination repair (HRR) genes, including BRCA1, BRCA2, ATM, BARD1, BRIP1, CHEK1, CHEK2, FAM175A, MRE11A, NBN, PALB2, RAD51C, and RAD51D (Pennington et al., 2014). Another study estimated that approximately 25.7% of ovarian cancer patients harbored germline or somatic mutations in one of the 16 HRR genes, including ATM, ATR, BARD1, BLM, BRCA1, BRCA2, BRIP1, CHEK2, MRE11A, NBN, PALB2, RAD51C, RAD51D, RBBP8, SLX4, and XRCC2 (Norquist et al., 2018).

Best characterized HRR biomarkers in high-grade serous ovarian cancer (HGSOC) are germline or somatic mutations in BRCA1/2 (Miller et al., 2020). Approximately 40% of ovarian cancer patients with family history of ovarian cancer can be attributed to BRCA1/2 mutations (Alsop et al., 2012). Women with BRCA1 and BRCA2 mutations have cumulative risk for developing ovarian cancer by age 80 as 44% and 17%, respectively (Kuchenbaecker et al., 2017). Frequency of BRCA1/2 mutations in HG-SOC was approximately 20.3%, dissected into approximately 14.6% germline and 6.0% somatic mutations (The Cancer Genome Atlas Research Network, 2011). Apart from BRCA1/2 mutations, additional 10.8% patients harbored BRCA1 promoter methylation (The Cancer Genome Atlas Research Network, 2011). Approximately 81% of BRCA1 mutations and 72.4% of BRCA2 mutations in HGSOC were accompanied by heterozygous copy-number loss, which suggested bi-allelic loss-of-function (BILOF) (The Cancer Genome Atlas Research Network, 2011).

In addition to BRCA1/2, deficiencies in non-BRCA1/2 HRR proteins, including RAD51, RAD54, DSS1, RPA1, NBS1, ATR, ATM, CHK1, CHK2, FANCD2, FANCA, and FANCC, have been shown to enhance poly(adenosine diphosphate–ribose) polymerase inhibitor (PARPi) sensitivity (McCabe et al., 2006). Germline mutations in BRIP1 (1.36%), PALB2 (0.63%), RAD51C (0.57%), RAD51D (0.57%), and BARD1 (0.21%) were significantly more common in ovarian cancer patients compared to general population, and mutation frequencies were not associated with race, histologic subtype or disease site (Norquist et al., 2016). Patients with somatic non-BRCA1/2 HRR gene mutations derived similar progression-free survival and overall survival as patients with BRCA1/2 mutations under platinum chemotherapy, compared to patients without any HRR mutation (Norquist et al., 2018).

RAD51C and RAD51D are moderate ovarian cancer predisposition genes, which have been suggested to be offered alongside BRCA1/2 in genetic testing in ovarian cancer (Loveday et al., 2011; Meindl et al., 2010; Song et al., 2015). As summarized by Song et al., 2015, the frequency of germline RAD51D mutations in ovarian cancer estimated by different studies ranged from 0.34% to 1.1% (0.34%, 0.35%, 0.55%, 0.82%, 1.1%) (Song et al., 2015).

Retrospective biomarker analysis of ARIEL3 study showed that, germline/somatic mutations on BRCA1, BRCA2, RAD51C, and RAD51D were predictors of exceptional benefit (exceptional benefit: progression-free survival at least 2 years) from PARPi (O’Malley et al., 2022). Approximately 2.3% of patients harbored germline or somatic RAD51C or RAD51D mutation, and these patients had higher frequency of exceptional benefit compared to patients harboring mutations in other non-BRCA1/2 HRR genes (60% versus 5%, respectively) (O’Malley et al., 2022). Retrospective biomarker analysis of ARIEL2 study showed that germline/somatic RAD51C mutation, RAD51D mutation, and high-level BRCA1 promoter methylation were predictors of response to PARPi, with a similar strength to BRCA1/2 mutations (Swisher et al., 2021).

In this study, we sought to provide evidence supporting RAD51D as an important non-BRCA1/2 HRR gene in Chinese ovarian cancer patients, from perspectives including prevalence, correlation with HRD score, and correlation with efficacy.

## 2. Methods

This study is a post-hoc analysis of the first-line adjuvant chemotherapy (FACT) cohort of EOC-HRD study (Li et al., 2023). Eligibility criteria for FACT cohort included being at least 18 years old at diagnosis; epithelial ovarian cancer, fallopian tube cancer, or primary peritoneal cancer; high-grade serous or grade-3 endometroid histological type; FIGO stage II, III, or IV; surgery conducted; at least five rounds of chemotherapy administered; no maintenance therapy of any kind; no PARPi administered for any purpose; date of last dose of adjuvant chemotherapy between 2009/12/01 and 2020/05/01. Medical records of patients were retrospectively surveyed from Peking Union Medical College Hospital and West China Second University Hospital. Baseline clinical characteristics of FACT cohort patients are summarized in Table 1.

**Table 1:** Baseline clinical characteristics.

| Characteristic | Level | RAD51D<br>wild-type (n=369) | RAD51D<br>mutated (n=11) |
| --- | --- | --- | --- |
| Age, years |  | 53 (47-61) | 61 (52-63.5) |
| Cancer type | Fallopian tube cancer | 39 (10.6) | 3 (27.3) |
|  | Ovarian cancer | 328 (88.9) | 8 (72.7) |
|  | Primary peritoneal cancer | 2 (0.5) | 0 (0) |
| Histological type | Grade-3 endometrioid | 7 (1.9) | 1 (9.1) |
|  | High-grade serous | 362 (98.1) | 10 (90.9) |
| FIGO stage | II | 29 (7.9) | 4 (36.4) |
|  | III | 297 (80.5) | 5 (45.5) |
|  | IV | 43 (11.7) | 2 (18.2) |
| Surgery type | IDS | 184 (49.9) | 4 (36.4) |
|  | PDS | 185 (50.1) | 7 (63.6) |
| Surgery residual | R0 | 226 (61.2) | 7 (63.6) |
|  | R1 | 107 (29) | 0 (0) |
|  | R2 | 36 (9.8) | 4 (36.4) |
| Concurrent use of bevacizumab | Without | 340 (92.1) | 11 (100) |
|  | With | 29 (7.9) | 0 (0) |
| Round of chemotherapy |  | 8 (6-9) | 7 (6-8) |
| Pre-treatment CA125, U/ml |  | 712 (250.4-1544) | 353.6 (80.9-630.05) |

EOC-HRD study investigated 28 HRR genes, which constituted the HRR28 gene list. The HRR28 gene list included ATM, ATR, BAP1, BARD1, BRCA1, BRCA2, BRIP1, CDK12, CHEK1, CHEK2, EMSY, FAM175A, FANCA, FANCC, FANCD2, FANCI, FANCL, MRE11A, NBN, PALB2, PPP2R2A, PTEN, RAD50, RAD51B, RAD51C, RAD51D, RAD54B, and RAD54L. A patient was defined as HRR28 mutated if having at least one of the HRR28 genes mutated. Methodological details regarding homologous recombination deficiency (HRD) assay development and molecular analyses may refer to Li et al., 2023.

The efficacy endpoint of this study was progression-free survival (PFS), defined as the time from last dose of chemotherapy to disease progression or death, whichever occurred first. PFS was estimated using Kaplan–Meier method. Treatment effect differences were assessed using log-rank test. Hazard ratio (HR) and associated 95% confidence interval (CI) were calculated using CoxPH model, adjusted for age, FIGO stage, surgery residual, surgery type, pre-treatment CA125, concurrent use of bevacizumab, and the round of chemotherapy. HR and associated 95% CI for subgroup analyses were calculated using unadjusted CoxPH model. Statistical analyses were performed in R 4.2.1.

## 3. Results

### 3.1. Prevalence of RAD51D alterations

The EOC-HRD FACT cohort contained 380 patients, in whom 11/380 (2.9%) were RAD51D mutated (Figure 1a). Among the 11 RAD51D mutations, 8/11 (72.7%) were BILOF (Figure 1b). Figure 1a together with Figure 1c illustrate that BRCA1/2 mutation and RAD51D mutation had no overlap, in another word they were mutually exclusive in the personnel level. As we can see in Figure 1d, among all the mutations in one of the HRR28 genes, BRCA1 was the most prevalent and BRCA2 was following not too closely. RAD51D, which was mutated 11 times, was close to half of the 25 times that BRCA2 was mutated. Besides BRCA1/2 and RAD51D mutated patients, there were still 24/380 (6.3%) patients mutated in one of the remaining HRR28 genes (Figure 1c).

**Figure 1.**
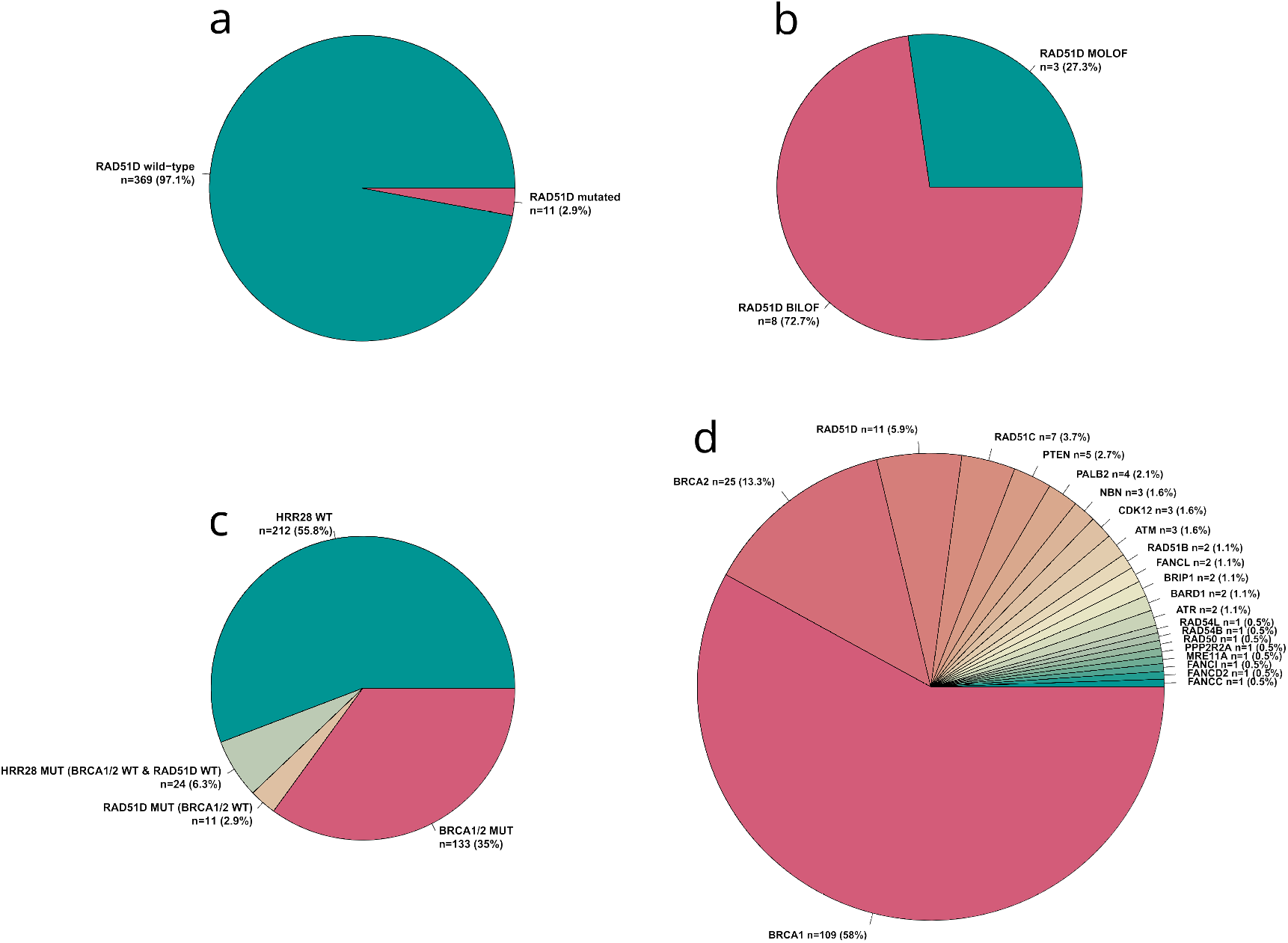
Mutation rate of HRR genes a) The frequency of RAD51D mutation was presented, and b) the proportion of MOLOF/BILOF within RAD51D mutations was further dissected. Frequency of HRR gene mutation was summarized c) by patient, and d) by HRR gene. HRR28 was a gene list composed of 28 HRR genes, as defined in the methods section. A patient was defined as HRR28 mutated if having at least one of the HRR28 genes mutated. MUT, mutated; WT, wild-type; MOLOF, mono-allelic loss-of-function; BILOF, bi-allelic loss-of-function.

### 3.2. Correlation with HRD score

As expected, BRCA1/2 mutation correlated well with HRD score, and BRCA1/2 BILOF had an even stronger correlation with HRD score (Figure 2a). When being used as the reference standard for training HRD score threshold, BRCA1/2 alteration is usually required to be BILOF (Li et al., 2023; Telli et al., 2016), implying that the correlation between BRCA1/2 mutation and HRD score won’t be as strong as the correlation between BRCA1/2 BILOF and HRD score. This is reasonable since BRCA1 and BRCA2 are both tumor suppressor genes, requiring both alleles to loss their functions in order to drive homologous recombination deficiency to the maximum capacity (Lewis, 2010). Compared with the RADA51D wild-type, RAD51D mutations elevated median HRD score from 56 to 65 with a p-value of 0.295, RAD51D BILOF elevated median HRD score from 56 to 65.5 with a p-value of 0.087 (Figure 2b). Besides BRCA1/2 and RAD51D, we also explored two additional widely recognized key HRR genes, RAD51C and PALB2 (Figure 2c-d). However, the alteration of neither RAD51C nor PALB2 had a positive correlation with HRD score. To sum up, although in term of prevalence rate, several non-BRCA1/2 HRR genes may come close to RAD51D, including RAD51C and PALB2; but in term of the correlation with HRD score, RAD51D was a clear winner compared to other non-BRCA1/2 HRR genes.

**Figure 2.**
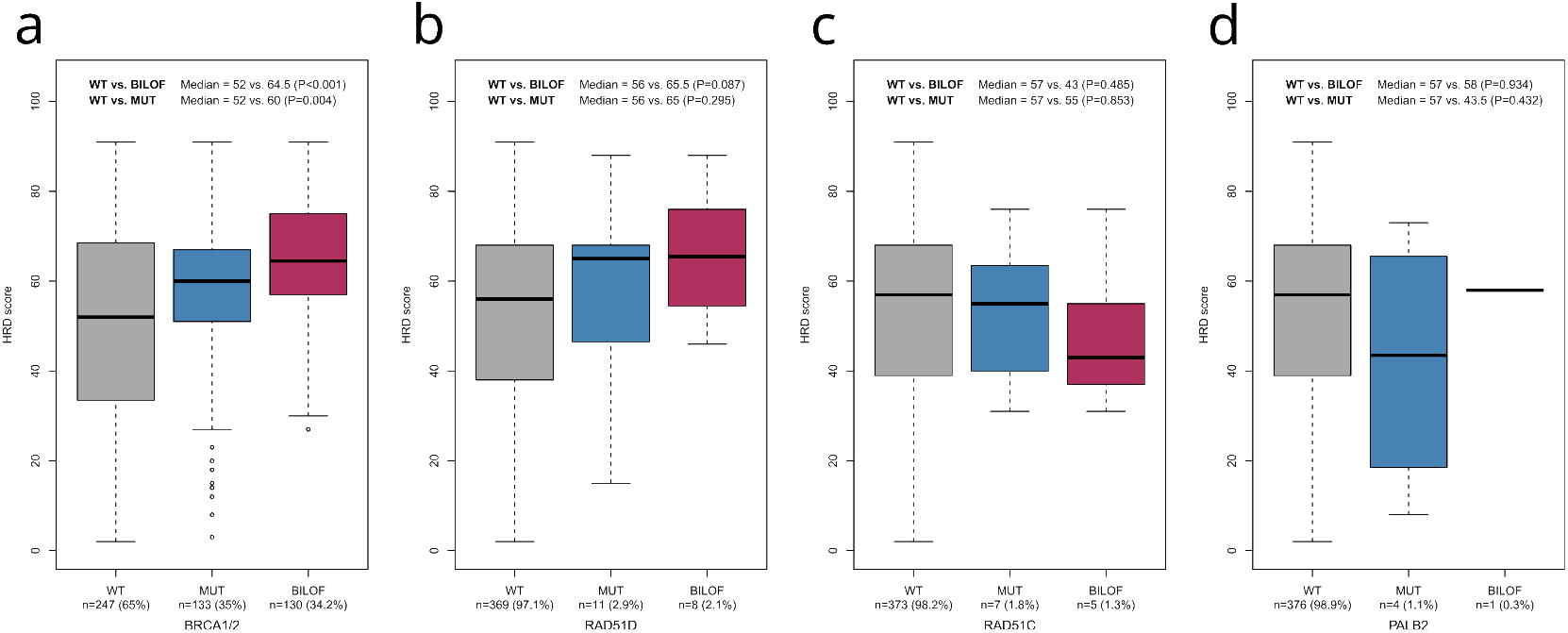
Correlation between HRR gene alteration and HRD score Boxplots visualizing HRD score distribution stratified by a) BRCA1/2 alteration, b) RAD51D alteration, c) RAD51C alteration, and d) PALB2 alteration. On each boxplot, the lower whisker, lower bound of box, center line, upper bound of box, and upper whisker represented the minimum, lower quartile, median, upper quartile, and maximum, respectively. P-values were calculated by the Wilcoxon test. MUT, mutated; WT, wild-type; BILOF, bi-allelic loss-of-function.

### 3.3. Correlation with efficacy

Figure 3a-b present that RAD51D mutation was a marginally significant predictor for PFS (median PFS increased from 13 to 25.2 months; HR 0.685; 95% CI, 0.314-1.494; P=0.069), RAD51D BILOF was a stronger but still marginally significant predictor for PFS (median PFS increased from 13 to 36.5 months; HR 0.571; 95% CI, 0.23-1.416; P=0.054). Since BRCA1/2 mutation is usually tested in routine clinical genetic testing for ovarian cancer patients, we should further evaluate how well RAD51D alteration correlates with PFS in the BRCA1/2 wild-type patients. Figure 3c-d present that, in the BRCA1/2 wild-type patients, RAD51D mutation was a statistically significant predictor for PFS (median PFS increased from 11.6 to 25.5 months; HR, 0.444; 95% CI 0.208-0.947; P=0.031), and RAD51D BILOF was an even stronger predictor for PFS (median PFS increased from 11.6 to 36.5 months; HR, 0.375; 95% CI 0.154-0.916; P=0.025). Therefore, regardless of being in the full population or the BRCA1/2 wildtype patients, RAD51D was a strong predictor for first-line chemotherapy efficacy.

**Figure 3.**
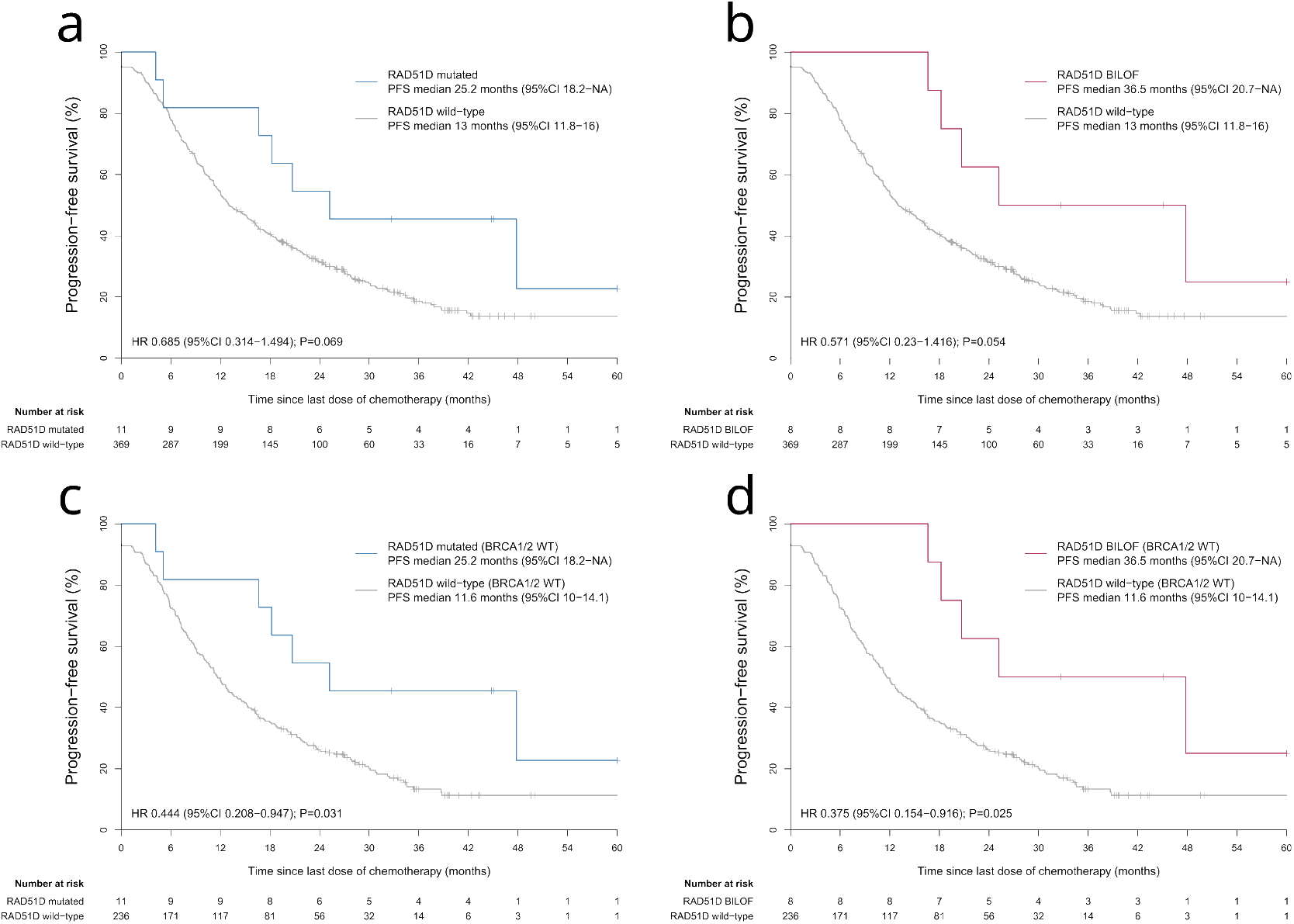
Correlation between RAD51D alteration and PFS The correlation between PFS and a) RAD51D mutation, b) RAD51D BILOF. The correlation between PFS and c) RAD51D mutation, d) RAD51D BILOF in the BRCA1/2 wild-type patients. WT, wild-type; BILOF, bi-allelic loss-of-function.

### 3.4. HRR3 gene list

A further question worth of being explored was whether combining RAD51D with BRCA1/2 to form a HRR3 gene list was worthy. HRR3 was defined as a gene list composed of 3 HRR genes (BRCA1, BRCA2 and RAD51D). A patient was defined as HRR3 mutated if having at least one of the HRR3 genes mutated. Before assessing the correlation between HRR3 mutation and PFS, we first took a look at the correlation between BRCA1/2 mutation and PFS as a benchmark. Figure 4a presents that, BRCA1/2 mutation was a statistically significant predictor for PFS (median PFS increased from 12.1 to 17.7 months; HR, 0.629; 95% CI, 0.487-0.814; P=0.003). Figure 4b presents that, compared with BRCA1/2 mutation, HRR3 mutation was a slightly stronger predictor for PFS (median PFS increased from 11.6 to 18.4 months; HR, 0.613; 95% CI, 0.476-0.788; P<0.001), and at the same time HRR3 had a higher mutation rate than BRCA1/2 (37.9% versus 35%).

**Figure 4.**
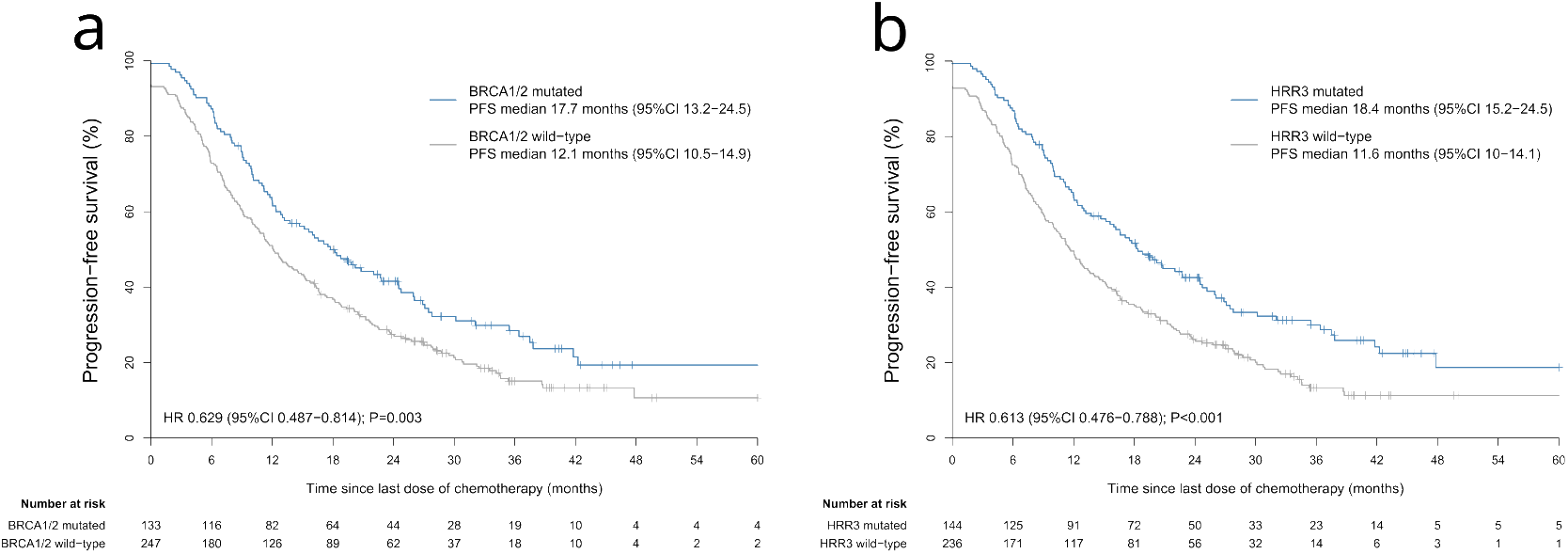
Correlation between HRR3 mutation and PFS The correlation between PFS and a) BRCA1/2 mutation, b) HRR3 mutation. HRR3 was a gene list composed of 3 HRR genes (BRCA1, BRCA2 and RAD51D). A patient was defined as HRR3 mutated if having at least one of the HRR3 genes mutated.

Data are median (IQR) or n (%). Surgery residual: R0, no residual; R1, residual < 1cm; R2, residual >= 1cm. Concurrent use of bevacizumab: with concurrent use of bevacizumab is defined as having received at least one dose of bevacizumab during firstline treatment. Round of chemotherapy: round of chemotherapy is the sum of round of first-line neoadjuvant chemotherapy and round of first-line adjuvant chemotherapy. Pre-treatment CA125: pre-treatment CA125 is the CA125 level measured before any form of first-line treatment is administered, including surgery and chemotherapy. Abbreviations: FIGO, International Federation of Gynecology and Obstetrics; PDS, primary debulking surgery; IDS, interval debulking surgery.

## 4. Discussion

Endogenous and exogenous sources of DNA damage continuously threaten genome stability, which may eventually give rise to cancer (Chatterjee & Walker, 2017; Grundy et al., 2020). DNA damage repair (DDR) mechanisms are vitally important in both cancer prevention and cancer treatment. HRR is a high-fidelity DDR pathway, with RAD51 being one of the key proteins in HRR pathway (Grundy et al., 2020). Utilities of RAD51 and its paralogs in cancer therapeutics can be either as biomarkers (prognostic, predictive, etc.) or as therapeutic targets. Tumors with deficient HRR system are sensitive to drugs interfering with DNA replication, but over-active HRR system may induce cancer treatment resistance or even metastasis (Broustas & Lieberman, 2014; Evers et al., 2010).

RAD51 and its paralogs function through complexes, including BCDX2 complex (RAD51B, RAD51C, RAD51D, and XRCC2), CX3 complex (RAD51C and XRCC3), RAD51C-RAD51-BRCA2-PALB2 complex, and Shu complex (SWSAP1 and SWS1) (Bonilla et al., 2020; Sullivan & Bernstein, 2018). RAD51 and its paralogs are ATPase with conserved Walker A and B motifs (Bonilla et al., 2020). Five of the six RAD51 paralogs are considered canonical paralogs (RAD51B, RAD51C, RAD51D, XRCC2, XRCC3), which share conserved amino acid sequences with RAD51, particularly around the Walker A and B motifs (Sullivan & Bernstein, 2018). The more recently discovered RAD51 paralog, SWSAP1, is a more divergent paralog (Sullivan & Bernstein, 2018). The most widely recognized RAD51 mediator is BRCA2, and other key mediators included PALB2, RAD54, plus the RAD51 paralogs (Bonilla et al., 2020; Grundy et al., 2020; Prakash et al., 2015). Disrupting RAD51 activity may be accomplished by either directing mutating RAD51 or through alterations of one of its mediators (Prakash et al., 2015; Van Der Zon et al., 2018).

Abnormalities on RAD51 activity or one of its mediators may give rise to cancers or Fanconi anemia (FA) (Prakash et al., 2015; Sullivan & Bernstein, 2018). FA is a rare genetic disease affecting mainly skeletal and immune systems, with symptoms including bone marrow failure, skeletal defects, premature aging, and elevated predisposition to other cancers (Moldovan & D’Andrea, 2009; Nalepa & Clapp, 2018). BILOF in one of the FA genes may give rise to FA (Nalepa & Clapp, 2018). FA genes include RAD51 (FANCR) and a number of its paralogs and mediators, including but not limited to RAD51C (FANCO), XRCC2 (FANCU), BRCA2 (FANCD1), and PALB2 (FANCN) (Nalepa & Clapp, 2018). Loss of RAD51 function may elevate the risk of cancer, whereas over-active RAD51 may cause cancer treatment resistance (Klein, 2008; Laurini et al., 2020). Maintaining an appropriate level of RAD51 activity is essential for HRR functionality, therefore essential for DDR, genome stability, and cancer prevention.

Functions of RAD51 and its paralogs mainly include DNA double-strand break (DSB) repair through HRR, and replication stress response (Grundy et al., 2020). DNA DSB is one of the most toxic forms of DNA damage (Khanna & Jackson, 2001), and can be repaired by HRR with high-fidelity (Ranjha et al., 2018). The high-fidelity of HRR originated from the fact that it uses homologous template for repair (Ranjha et al., 2018). After DNA DSB is formed, the following key RAD51-associated repairing procedures are conducted sequentially: DNA ends resected to form single-stranded DNA (ssDNA) overhangs; ssDNA coated by replication protein A (RPA); RPA displaced by RAD51 to form nucleoprotein filament (mediated by PALB2, BRCA2, BCDX2 complex, and CX3 complex); homology search and strand invasion by RAD51 filament to form D-loop structure (mediated by RAD54); RAD51 displaced to allow polymerases to copy from homologous template; second end of DSB captured; DSB repair intermediates resolved through resolution or dissolution (Bonilla et al., 2020; Grundy et al., 2020). The formation of RAD51 nucleoprotein filament is central to functions of RAD51 in DNA DSB repair through HRR (Grundy et al., 2020). On the other hand, RAD51 also has functions around stalled replication forks during DNA replication stress response, including but not limited to promoting replication fork reversal, protecting nascent strands of DNA from degradation by exonucleases, and restart of replication forks (Bonilla et al., 2020; Grundy et al., 2020).

RAD51D has a unique role that isn’t shared by other RAD51 paralogs, RAD51D serves in telomere maintenance (Tarsounas et al., 2004). Telomere erosion caused by RAD51D dysfunction might be a contributor to genome instability (Tarsounas et al., 2004). Secondary somatic reversion mutations on RAD51D have been denoted as an acquired PARPi resistance mechanism, therefore supporting the role of RAD51D in synthetic lethality with PARPi (Kondrashova et al., 2017).

We observed in this study that, in term of correlation with HRD score and correlation with efficacy, RAD51D BILOF had greater strength then RAD51D mutation (Figure 2, 3). This phenomenon could be explained by the fact that RAD51D is a tumor suppressor gene, and alterations of tumor suppressor genes tend to be recessive in nature (Lewis, 2010). Therefore, RAD51D likely need both alleles to loss their functions in order to drive homologous recombination deficiency to its maximum capacity.

The major limitation of this study is that all analyses originated from a retrospective study with two study centers, so we cannot rule out that parts of our findings might be prone to randomness. Therefore, we are looking forward to future studies for validating the findings of this study.

Ethier et al., 2022 assessed the potential predictive DDR biomarkers for patient selection or stratification in clinical studies in epithelial ovarian cancer. BRCA1/2 mutation and HRD status were assigned level-of-evidence (LOE) I. RAD51C/D mutation, along with BRCA1/2 reversion mutation, BRCA1 methylation, and CCNE1 amplification, were assigned LOE II, therefore awaiting more clinical validations (Ethier et al., 2022).

In this study, we provided our piece of evidence supporting RAD51D as an important non-BRCA1/2 HRR gene in Chinese ovarian cancer patients, from perspectives including prevalence, correlation with HRD score, and correlation with efficacy.

## 5. Acknowledgement

## 5.1. Ethics committee approval and informed consent

This study is a post-hoc analysis of the first-line adjuvant chemotherapy cohort of EOC-HRD study (Li et al., 2023). EOC-HRD study was approved by the Ethics Committee of Peking Union Medical College Hospital, Beijing (project ID: JS–1932), written informed consent was obtained from every patient.

## 5.2. Funding statement

This study is a post-hoc analysis of the first-line adjuvant chemotherapy cohort of EOC-HRD study (Li et al., 2023).

EOC-HRD study was supported by a grant from CAMS Innovation Fund for Medical Sciences (CIFMS-2017-I2M-1–002), and grants from the National High Level Hospital Clinical Research Funding (No. 2022-PUMCH-A-117, 2022-PUMCH-B-083, 2022-PUMCH-C-010, 2022-PUMCH-C-022, and 2022-PUMCH-D-003), and the Le Fund (No. KH-2020-LJJ-004, 034, and 035).

This study was supported by grants from: 1. The Key Project of Sichuan Provincial Department of Science and Technology (2019YFS0532) (Ethical Lot Number: 20220129);

2. Horizontal Science and Technology Project of Sichuan University (23H1223); 3. The Project of Chengdu Science and Technology Administration (2021-YF05-01725-SN).

## 5.4. Data availability

Raw sequencing data of EOC-HRD study patients cannot be publicly shared as patients did not consent to share raw sequencing data beyond the research and clinical terms. The datasets generated and analyzed might be available upon reasonable request and data usage agreement.

